# Machine Learning Driven ‘Therapy Calculator’ for Self-Managed Digital Speech-Language Therapy for Individuals with Post-stroke Aphasia

**DOI:** 10.64898/2026.01.22.26344656

**Authors:** Hantian Liu, Margrit Betke, Prakash Ishwar, Swathi Kiran

## Abstract

Individuals with post-stroke aphasia live with long-term disabilities, yet they do not know whether they will improve their communication and cognitive skills over time. We propose a “Therapy Calculator” to provide patients with a better understanding of likely recovery as they engage with therapy. Using a large dataset of rehabilitation outcomes from a digital therapeutic called Constant Therapy (3.5 million therapy sessions of 18,000+ users), we developed a machine learning algorithm that estimates the probability of improvement from one functional landmark (i.e., a given skill level) to the next in a functional domain (e.g., reading) while accounting for age, etiology, starting performance, and frequency and duration of therapy. This logistic regression model performed a binary classification task, i.e., whether patients can improve to the next landmark, with an average F1 score of all models at 0.84, suggesting reliable prediction of moving to the next landmark. Then, we created an online “Therapy Calculator” to assess a new user’s current functional level and demographic information, and make predictions by passing these features into models trained on relevant subsets of historical data. The findings indicate that our model can provide reliable predictions for patients beginning self-managed SLT, and therapy calculator is publicly available.

## Introduction

Speech-language therapy (SLT) provides benefits for the functional use of language in patients with aphasia^1^. Practically, however, the amount of SLT received by patients in a clinical setting is highly limited by multiple factors including constraints on insurance and reimbursements, difficulties with mobility and travel, geographic isolation, etc. Although previous studies have shown that an average total of 98.4 hours of therapy can lead to positive outcomes^2^, typically stroke survivors only receive 8 hours of SLT in the year after their stroke^3^.

To address the issue of insufficient SLT dosage, computer-based therapy approaches have enabled high-intensity, self-managed practice, yielding significant language improvements^4–7^. For instance, Liu *et al*.^6^ demonstrated that consistent, long-term use of digital therapeutics for language and cognitive deficits led to measurable and sustained gains over a span of 30 weeks. Fleming *et al*.^8^ showed significant improvements in auditory comprehension using a self-managed app for chronic post-stroke aphasia, while the BIG CACTUS randomized control trial confirmed the efficacy of intensive computer-based naming treatment in post-stroke aphasia patients^9^. Despite these benefits, Nichol *et al*.^7^ have highlighted barriers such as limited digital literacy and variable tech support, thereby emphasizing the need for SLTs to provide tailored guidance to optimize outcomes. Nonetheless, emerging clinical evidence underscores the effectiveness of structured, computer-based therapies in improving communication outcomes^10^.

One such self-managed digital therapy application is called Constant Therapy, a commercially available digital therapeutic software. Several studies have demonstrated the clinical efficacy of Constant Therapy (CT) software for language and cognitive rehabilitation. For instance, the RecoverNow trial highlighted its feasibility for acute post-stroke aphasia, with 83% of patients engaging in daily app-based practice for an average of 60 minutes during hospitalization^5^. Edgar and Bargmann^11^ emphasized CT’s adaptability and effectiveness in both traditional and telepractice settings for individuals with dementia. Using the large database of user data collected within the application, Cordella *et al*.^12^ conducted a retrospective analysis linking higher therapy dosage (practice upto 4 or 5 times per week) to greater outcomes. A similar retrospective analysis showed that at-home CT users practiced more frequently and achieved faster progress compared to in-clinic users^12,13^. In addition, Braley *et al*. (2021)^14^ conducted a 10-week pilot randomized control trial with 32 post-stroke aphasia patients, comparing self-managed CT therapy with workbook therapy. The CT group demonstrated significantly greater improvements than the workbook group in WAB-R Aphasia Quotient (mean change: 6.75 vs. 0.38), surpassing the 5-point clinically significant benchmark^15^. High compliance with the prescribed regimen (30 minutes, five days per week) highlighted the feasibility of this approach. These studies support the efficacy of Constant Therapy as a self-managed digital therapy for post-stroke aphasia and further underline the importance of consistent therapy practice for optimal rehabilitation outcomes.

In computer-based treatments that can be self-managed (i.e., patients log in and out of a therapy practice session), patients can control their therapy schedule, including practice time and frequency. In most cases, however, they are not aware of how much therapy / practice they would need to achieve in order to see improved outcomes. With the development of machine learning techniques, it is possible to make such a prediction based on performance of past patients to provide estimates of required therapy practice for new patients. The goal of this study is to develop a therapy practice calculator that can first assess the functional level of new patients, and, based on data-driven assumptions, predict the probability of them improving in their therapy outcomes based on specific practice frequency and total practice time.

To achieve these goals, the present study developed an application, called Therapy Calculator, that includes a machine learning (ML) based predictor and a user-friendly web interface (Figure 1). An ML algorithm was trained based on available data from previous patients who used Constant Therapy (details below). The Therapy Calculator first gathers basic demographic information from new patients and presents them with a short assessment process to estimate their current functional level within a language or cognitive domain selected by the patient. It then asks the patients to select the frequency and practice length of their planned therapy using a slider interface, and runs the ML algorithm to predict the likelihood of the patients’ improvement from the initial performance based on these selections. The sliders enable the patients to change their original practice schedule easily in order to see whether or by how much additional therapy would increase the likelihood of their improvement. The Therapy Calculator is accessible to users as a web-based app. Its is freely available at https://www.bu.edu/cbr/research/predictive-modeling/ In this paper, we describe the (1) development and evaluation of the ML algorithm to predict the probability of performance improvement and (2) the development and deployment of the Web-based Therapy Calculator.

**Figure 1.**
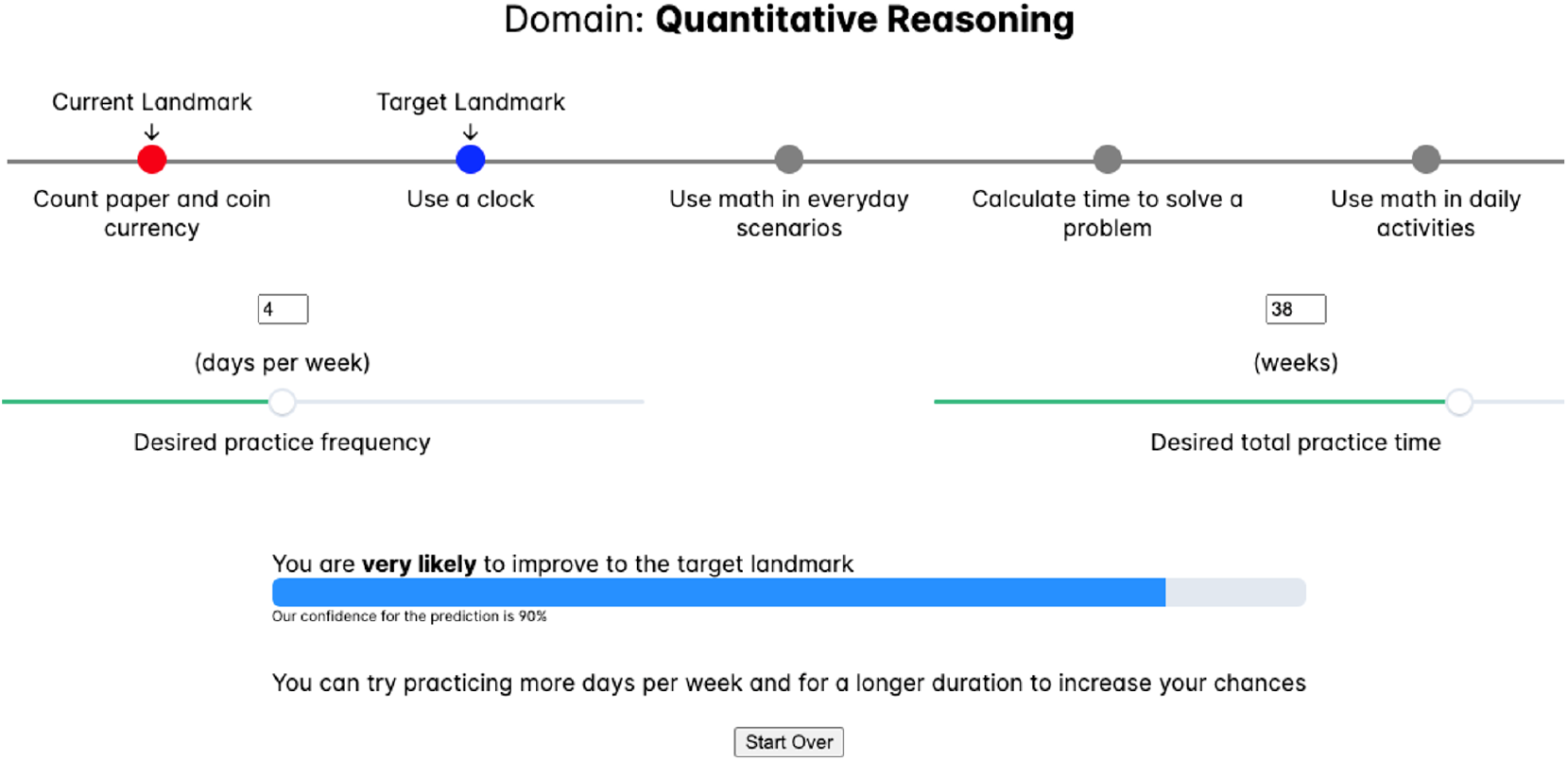
The Therapy Calculator predicts that a specific patient who plans to work on quantitative reasoning tasks 4 days a week for 38 weeks will very likely improve from the current (first) to the target (second) landmark.

## Methods

### Constant Therapy Program

Constant Therapy is a speech, language, and cognitive therapy app that allows digital therapy practice (e.g., picture naming, sentence completion, math problems, memory exercises, categorization tasks, reading comprehension exercises, or visuospatial puzzles) following a self-managed schedule. The tasks on the app are organized into various functional domains. This study focused on 13 domains: (1) analytical, (2) arithmetic, (3) attention, (4) auditory comprehension, (5) auditory memory, (6) naming, (7) phonological processing, (8) production, (9) quantitative, (10) reading, (11) visual memory, (12) visuospatial skills, and (13) writing. Each domain comprises multiple tasks arranged in order of increasing difficulty specifying the *task progression order*. Patients start at a certain level and move to a higher-or lower-difficulty task based on their performance. The progression from task-to-task is guided by the app’s adaptive learning algorithm that adjusts the difficulty based on real-time performance data^16,17^. Each task on the app is designed to target specific therapeutic goals. In this way, each patient receives personalized therapy tasks, enabling them and their clinicians to identify high-priority, functionally-relevant therapy goals^18^.

To calculate improvements on the task, the app uses a domain score to evaluate the patient’s progression through tasks within a domain. It is a numeric value ranging from 0 to 1, where 0 indicates no progress in the domain and 1 represents full completion of tasks within the domain^19^. To aid the interpretibility of domain score, functional landmarks are employed to contextualize these scores into discernible abilities within each domain. For example, in the auditory comprehension domain, landmarks might include understanding words (Level 0), understanding sentences (Level 1), understanding a short story (Level 2), and inferring information from long messages (Level 3), representing a progression from simpler to more complex cognitive abilities. At each landmark for each patient, the landmark score indicates whether the patient has achieved the functional ability by having a domain score more than the landmark score. For example, the landmark scores for each of the landmarks above are 0, 0.38, 0.81, and 1 respectively.

### Data Filtering

For this study, we used a dataset provided by Constant Therapy that is described and used in previous studies^6,19^. The dataset contains 3,556,321 therapy session records from 18,728 unique patients, collected between April 2020 and June 2023. To ensure the relevance and quality of the data, we applied various filters to both the therapy records and the patient data. A flowchart of the filtering process is shown in Figure 2. The first stage selects patients that actually subscribed to the app and were assigned practice schedules by the system. This ensures sufficient time of engagement and excludes casual users. It also includes patients who completed their assessments independently, i.e., without the assistance/supervision of a speech pathologist. As previously noted, we focused on self-managed patients who practiced therapy independently, however, since clinicians may also use the application to help with their patients’ recovery process, such data were filtered and removed from the data analysis. Furthermore, to select the most active and continuously engaged users, we further filtered the data based on the following inclusion criteria: (1) patients must have more than 10 sessions in a given domain, (2) the maximum time gap between two consecutive sessions must be less than 14 days, and (3) patients must have age and time since stroke information recorded in the system. The data filtering process yielded records of 2,269 patients, which we used for training our ML algorithm as described in the next section.

**Figure 2.**
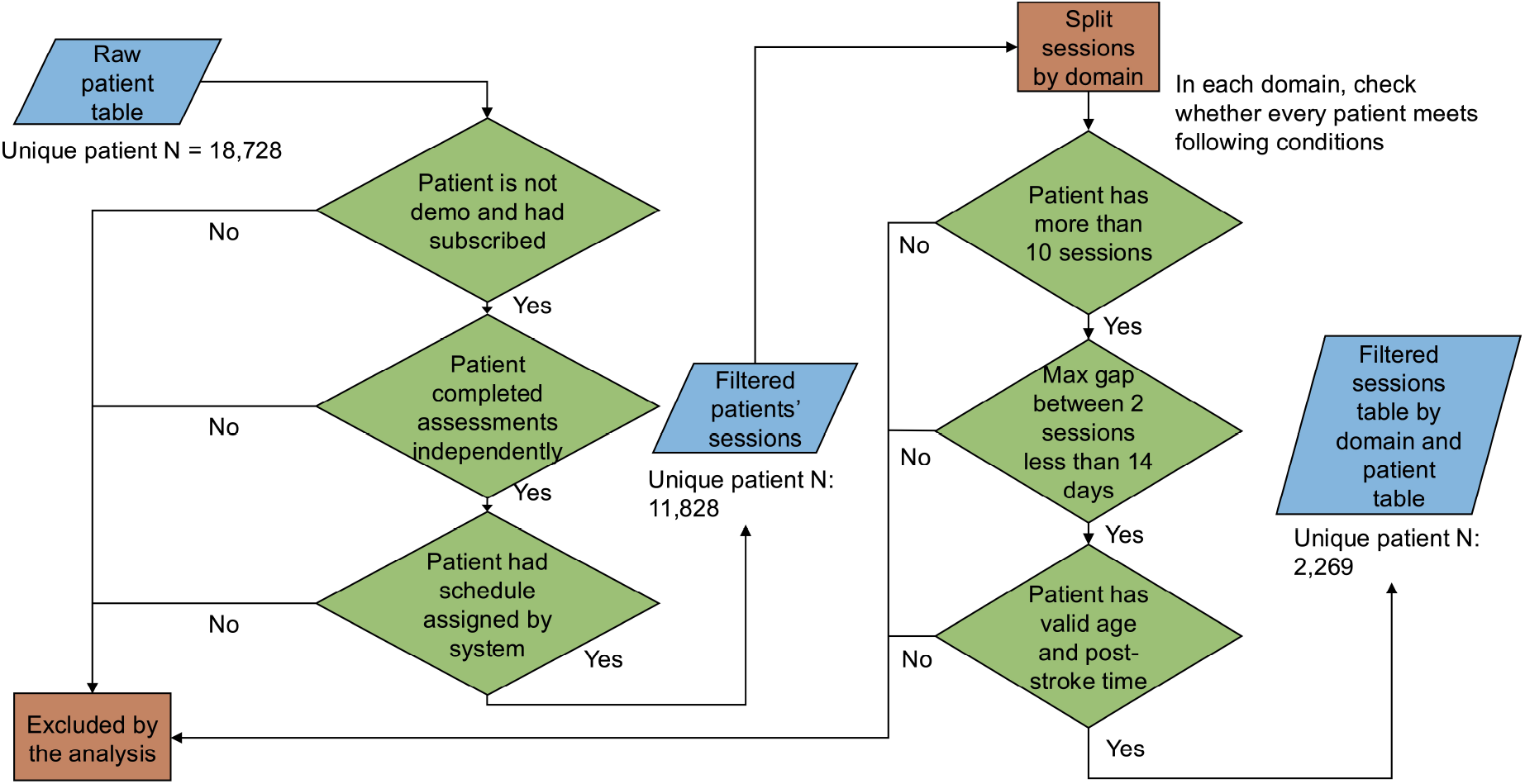
A flowchart of data filtering process. In the first stage of filtering, only self-managed patients are selected. In the second stage of filtering, only patients that have a minimum level of activity and continuous engagement are selected.

### Machine Learning Approach for Therapy Calculator

After filtering patient data, we selected features that captured both the current functional level and the anticipated therapy engagement patterns of these patients. These are: (1) age of the patient in years; (2) time since stroke in months; (3) initial domain score; (4) initial task accuracy in percentage; (5) total number of therapy sessions within the domain; (6) total length of therapy in days; and (7) maximum time gap between two sessions in days. The first two demographic features are essential for personalized prediction. Features (3) and (4) are based on a subjective and an objective assessment administered to each user (see section on Web-based Interface for Therapy Calculator). They act as direct indicators of current abilities, providing the baseline measure for a given task/domain. The last three features indicate the dosage, duration and consistency of therapy practice, all of which potentially influence the rate and extent of recovery. Patients were grouped by their initial landmark based on their initial domain score, with the goal of predicting whether they will improve their domain score to the threshold of the next landmark during their therapy process. A positive label represents performance improvement and a negative label represents absence of performance improvement. This grouping facilitated targeted predictions that were specifically tailored to each individual patient’s starting level.

We selected logistic regression algorithm for our machine learning model due to its robustness and interpretability – a vital aspect when providing feedback to patients, while providing satisfactory prediction performance. Logistic regression uses the logistic model to calculate the probability of a patient reaching the next functional milestone. Specifically, suppose **x** = (*x*_1_, *x*_2_, …, *x*_7_) denotes the 7 features described in the previous paragraph after data-standardization (subtracting from each feature its mean value in the training dataset and dividing the result by the standard deviation of the feature in the training dataset – this makes each standardized feature have zero-mean and unit variance in the training dataset). The logistic model calculates the probability that a patient with standardized features given by **x** will reach the next functional milestone, i.e., have a positive label *y* = 1, as:

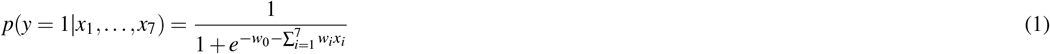

The probability that the patient will not reach the next functional milestone, i.e., *y* = 0, is 1 − *p*(*y* = 1 | *x*_1_, …, *x*_7_). The 8 parameters *w*_0_, *w*_1_, …, *w*_7_ of the logistic model are learned from the labeled training data by maximizing the overall probability (according to the logistic model) of all (**x**, *y*) pairs in the training data, where **x** are the standardized features of a patient and *y* the ground truth binary label for the patient. Since the features are standardized and influence the probability of reaching the next functional milestone through their linear combination, the magnitude and sign of coefficients *w*_1_, …, *w*_7_ can be used to interpret the impact of each feature (see next section).

For the training process we used a k-fold cross-validation technique to ensure generalizability. Specifically, data in each domain-landmark grouping were standardized, i.e., scaled to have zero mean and unit variance, and then randomly split into five subsets. In each training iteration, one of the five subsets served as a validation set while the remaining four were used to train the model. The aggregate performance of the model on each landmark was quantified by the mean of the validation scores obtained across the five folds. For model evaluation, we used F1 score as the primary metric, which provides a balance between precision and recall. Since many landmarks have an unbalanced number of samples in each class, accuracy is not the best metric to evaluate model performance^20^. Figure 3 describes the basic workflow of the training process.

**Figure 3.**
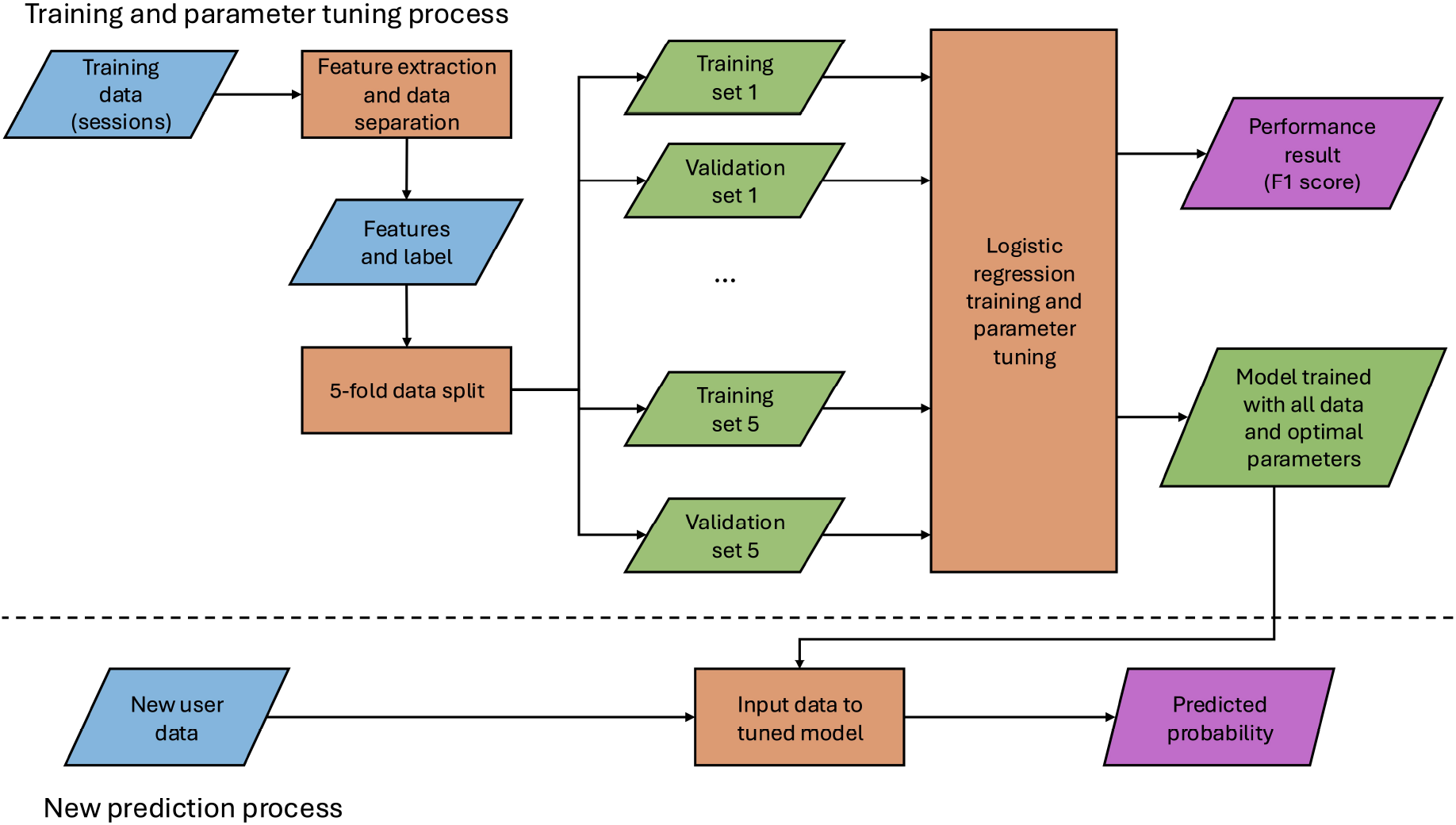
Flowchart of the training process

## Results of Model Prediction

The logistic regression algorithm achieved an average F1 score of 0.84 across all domain-landmark combinations, with a standard deviation of 0.10. We observed that lower-numbered landmarks, representing easier tasks within a domain, demonstrated higher F1 score. For example, the F1 score for the first landmark over all domains is 0.89, with 0.06 standard deviation, while for the highest landmark available in each domain, the F1 score is 0.81 over all domains and standard deviation is 0.10. This indicates that predictions for easier/earlier tasks (lower landmarks) within a domain are more reliable.

A heatmap of the F1 scores are shown in Figure 4 for each individual model for each landmark. The figure also shows the number of positive (patients who progressed to next landmark) and negative (patients who did not progress to next landmark) samples across each domain-landmark grouping. Note that we restricted model training to landmarks with more than 50 samples to ensure statistical reliability. Landmarks without a sufficient number of samples to make predictions are denoted as ‘N/A’ in the figure.

**Figure 4.**
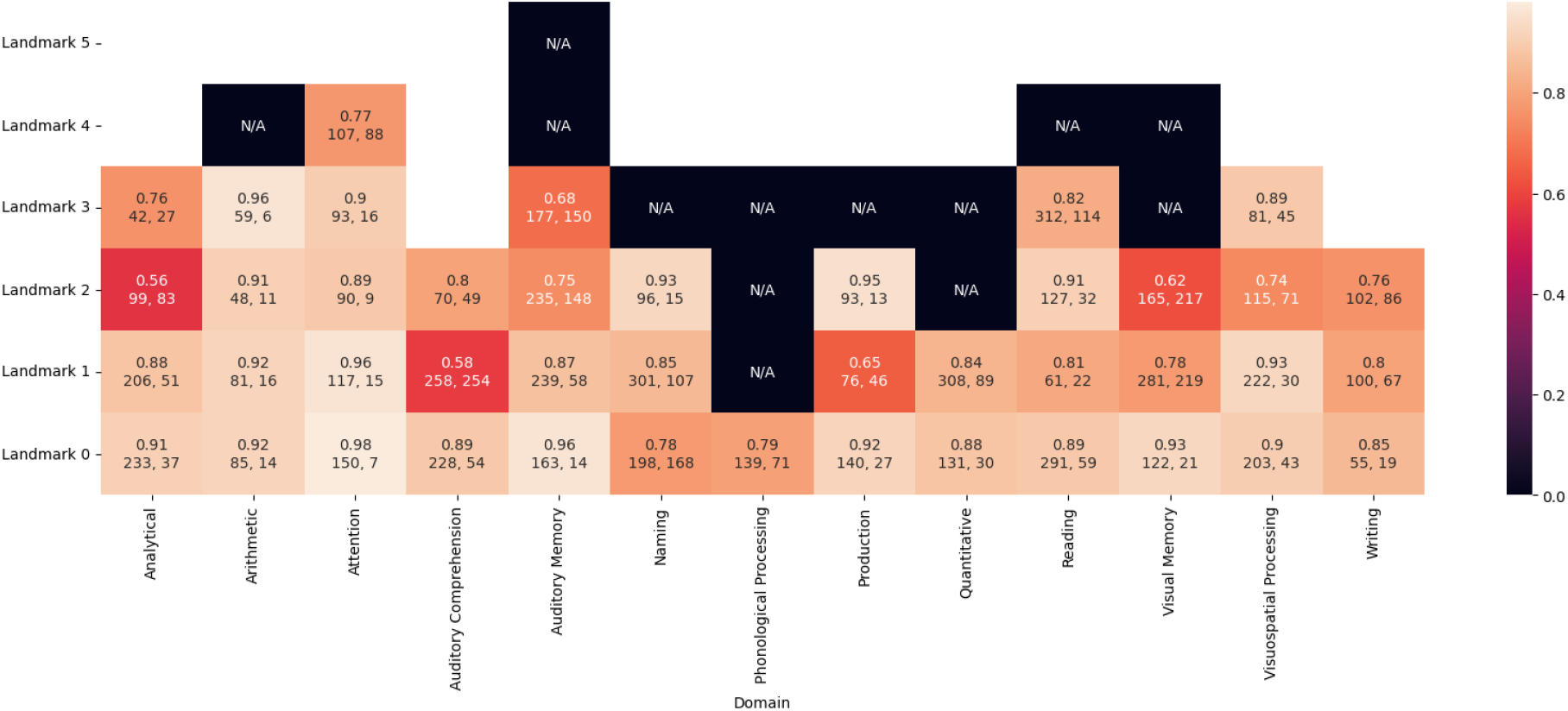
F1 scores and number of positive and negative samples for each landmark and each domain. Landmarks without a sufficient number of samples to make predictions are denoted as ‘N/A’. First row in each cell: F1 score of the landmark.

As described in Eq. 1, each input feature *x*_*i*_ has a corresponding weight *w*_*i*_. While all features are standardized to zero mean and one variance, evaluating the weight values provides critical insights regarding feature importance and its impact on the prediction outcomes. Larger absolute value of weights represents higher importance and the sign of the value indicates positive or negative impact on model performance. For each domain and for each of the seven features, we analyzed the weights for each model Table 1 shows the mean and standard deviation of model coefficients averaged over all landmarks within each domain.

**Table 1.** For each domain and all domains averaged (last row), the mean and standard deviation (indicated within parenthesis) of the model coefficient for each feature (i.e., *w*_*i*_) is shown. The type (T) of each domain is either cognitive (C) or speech/language (SL). The domain Phonological Processing contains only one landmark with enough data.

| Domain | T | Age | Time since stroke | Init domain score | Init accuracy | Total sessions | Therapy length | Max gap |
| --- | --- | --- | --- | --- | --- | --- | --- | --- |
| Analytical | C | -0.08 (0.32) | -0.002 (0.02) | 0.35 (0.47) | 0.58 (0.44) | 0.17 (0.24) | -0.47 (0.30) | 0.18 (0.22) |
| Arithmetic | C | -0.38 (0.29) | -0.24 (0.11) | 0.06 (0.13) | 0.17 (0.26) | 0.22 (0.23) | 0.23 (0.30) | 0.05 (0.22) |
| Attention | C | -0.01 (0.19) | -0.07 (0.40) | 0.39 (0.24) | 0.69 (0.18) | 0.45 (0.44) | 0.29 (0.79) | 0.13 (0.29) |
| Auditory Comprehension | SL | -0.19 (0.24) | -0.13 (0.03) | 0.39 (0.23) | 0.21 (0.08) | 0.02 (0.28) | 0.06 (0.18) | 0.08 (0.11) |
| Auditory Memory | C | -0.24 (0.22) | 0.01 (0.43) | 0.33 (0.29) | 0.23 (0.39) | 0.03 (0.34) | 0.52 (0.34) | 0.08 (0.36) |
| Naming | SL | -0.13 (0.08) | -0.17 (0.09) | 0.16 (0.32) | 0.58 (0.32) | 0.83 (0.98) | -0.02 (0.08) | -0.05 (0.17) |
| Phonological Processing | SL | -0.23 (N/A) | -0.26 (N/A) | -0.08 (N/A) | 0.83 (N/A) | -0.04 (N/A) | 0.26 (N/A) | -0.38 (N/A) |
| Production | SL | -0.03 (0.31) | 0.02 (0.08) | 0.67 (0.40) | 0.37 (0.26) | 0.29 (0.52) | 0.20 (0.48) | -0.19 (0.10) |
| Quantitative | C | -0.05 (0.23) | -0.18 (0.18) | 0.59 (0.15) | 0.16 (0.37) | 0.59 (1.16) | 0.24 (0.21) | -0.31 (0.06) |
| Reading | SL | -0.03 (0.28) | -0.22 (0.27) | 0.38 (0.25) | 0.37 (0.22) | 0.32 (0.28) | 0.44 (0.46) | 0.01 (0.21) |
| Visual Memory | C | -0.53 (0.39) | -0.35 (0.20) | 0.32 (0.38) | 0.53 (0.32) | 0.28 (0.74) | 0.71 (0.49) | -0.14 (0.32) |
| Visuospatial Processing | C | -0.03 (0.05) | -0.25 (0.10) | 0.71 (0.11) | 0.43 (0.38) | 0.64 (0.11) | -0.17 (0.48) | 0.05 (0.37) |
| Writing | SL | -0.30 (0.13) | -0.03 (0.33) | 0.23 (0.44) | 0.90 (0.25) | 0.28 (0.47) | -0.30 (0.49) | -0.11 (0.28) |
| All domains averaged |  | -0.17 (0.27) | -0.14 (0.24) | 0.37 (0.32) | 0.45 (0.34) | 0.33 (0.51) | 0.16 (0.51) | -0.02 (0.26) |

Among demographic factors, ‘Age’ has a consistently negative average coefficient value across all domains with an overall mean of −0.17 mean and an overall standard deviation of 0.27 (last row, third column of Table 1). The impact of age becomes more pronounced at higher-numbered landmarks (see supplementary table S1). This aligns with clinical observations that younger patients tend to experience more significant improvements in challenging tasks. The ‘Time since stroke’ feature also has a consistently negative (but smaller) influence across most domains with an overall mean coefficient value of −0.14 and an overall standard deviation of 0.24 (last row, 4th column of Table 1). This suggests that more acute patients have a higher likelihood of improving to the next landmark.

We now examine how patients’ current performance ability plays an important role in predicting therapy outcome. The ‘Initial domain score’ has a high positive influence across almost all domains and landmarks with an overall mean of 0.37 and an overall standard deviation of 0.32 (last row, column 5, in Table 1). Among all the 45 domain-landmark combinations, the initial domain score has a positive impact in 38 of them (supplementary table S1). That higher initial domain scores are a strong predictor of progression, suggests that less severe impairments are more likely to exhibit more improvements over time. ‘Initial task accuracy’ from the assessment phase has a positive impact in 42 domain-landmark combinations (supplementary table S1) with a high positive coefficient for mid-to high-numbered landmarks. In fact, ‘Initial task accuracy’ had the highest positive coefficient among all features. Both features indicate that better initial performance ability leads to better therapy outcomes.

Lastly, among features related to therapy dosage, ‘Total number of therapy sessions’ was one of the most influential features (0.33 mean, 0.51 standard deviation). The positive coefficients indicated that a higher frequency of scheduled therapy sessions strongly correlated with improvement across landmarks, particularly in domains requiring consistent practice, such as auditory comprehension and reading. The total length of therapy was surprisingly not a clear predictor; the coefficient for this feature showed considerable variation across landmarks, with negative coefficients for lower landmark values, but becoming less impactful for higher landmarks. Lastly, ‘max time gap’ between sessions exhibited negative coefficients for most models, reinforcing the importance of maintaining consistent practice intervals for optimal progress.

In addition to the interpretability of the feature-wise coefficients, it is also valuable to evaluate the impact of features in each domain. As mentioned above, the 13 domains analyzed in the study were categorized as cognitive domains and speech and Second row in each cell: left: number of positive samples, right: number of negative samples. language domains. Seven cognitive domains (type C domains in Table 1) showed strong dependency on demographic features, such as age and time since stroke. Younger patients in these domains tended to progress faster, particularly in tasks requiring quick adaptability, for example, attention and visuospatial skills domains. Patients with less time since injury exhibited higher improvement rates, especially in analytical and memory-related domains, reflecting the importance of early intervention for recovery. Smaller gap between sessions and shorter length of therapy yielded better results in most of the domains, indicating sustained engagement strengthens recovery and improvement, and patients who are able to improve tended to progress in relatively shorter time period. Tasks involving analytical or quantitative reasoning displayed slower progression for patients with longer gaps between sessions, likely due to the higher cognitive load these domains require. Initial task scores were moderately influential, suggesting that baseline abilities predict recovery, but not as strongly as in speech and language domains.

For speech and language domains (six type SL domains in Table 1), while less influential in lower landmarks, age significantly impacted higher-numbered landmarks in domains like reading and production. Older patients faced more difficulty in advancing, likely due to reduced motor coordination and slower processing speeds. Similar to cognitive domains, patients with lower time since injury displayed faster improvement in naming and auditory comprehension, where earlier/easier tasks correlated with better long-term outcomes. In these domains, initial task score was the most predictive feature, especially for phonological processing and writing, indicating that higher baseline performance strongly correlated with further improvement. Daily or nearly-daily practice was also essential for progress in these domains, underscoring the need for intensive engagement. Domains like writing and phonological processing were particularly sensitive to therapy frequency, as motor-skill-based recovery relies heavily on repetition and routine. The auditory comprehension domain was unique in its balanced reliance on both demographic and therapy dosage-specific factors, highlighting the need for a multifaceted therapy plan.

### Web-based Interface for Therapy Calculator and User Feedback

Once the model development and evaluation was completed with an estimated probability of new patients improving with therapy, the second goal of the project was to develop a web-based interface for convenient access to this calculator. The website was developed using Vue.js and Python, and deployed on Amazon Web Service (AWS). Specifically, the frontend was saved in AWS S3 and deployed on Cloudfront, and the backend was developed as Lambda functions that can be called via AWS API Gateway.

The workflow of the web-based interface of the Therapy Calculator for user input and front- and backend services is shown in Figure 5. The user enters basic demographic information such as age and time since injury. They are then asked to select the domain they would like the therapy calculator to make predictions for. Based on the selected domain, they are asked to complete two assessment tasks for the calculator to determine their current functional level within the domain. First, the frontend service presents questions that require patients to *subjectively* estimate their functional levels. Patients are asked how well they can perform daily tasks related to the domain and the answers range from *not at all* to *extremely well*. Based on their responses, the backend service fetches “objective assessment tasks” from the Constant Therapy server. Objective assessment tasks are sets of in-app tasks that aim to quantify the patient’s skill level at a certain level of task progression in the domain. Based on the accuracy of completing the objective assessment task, the frontend service assigns the user’s initial domain score, which represents the user’s current functional level in the domain.

**Figure 5.**
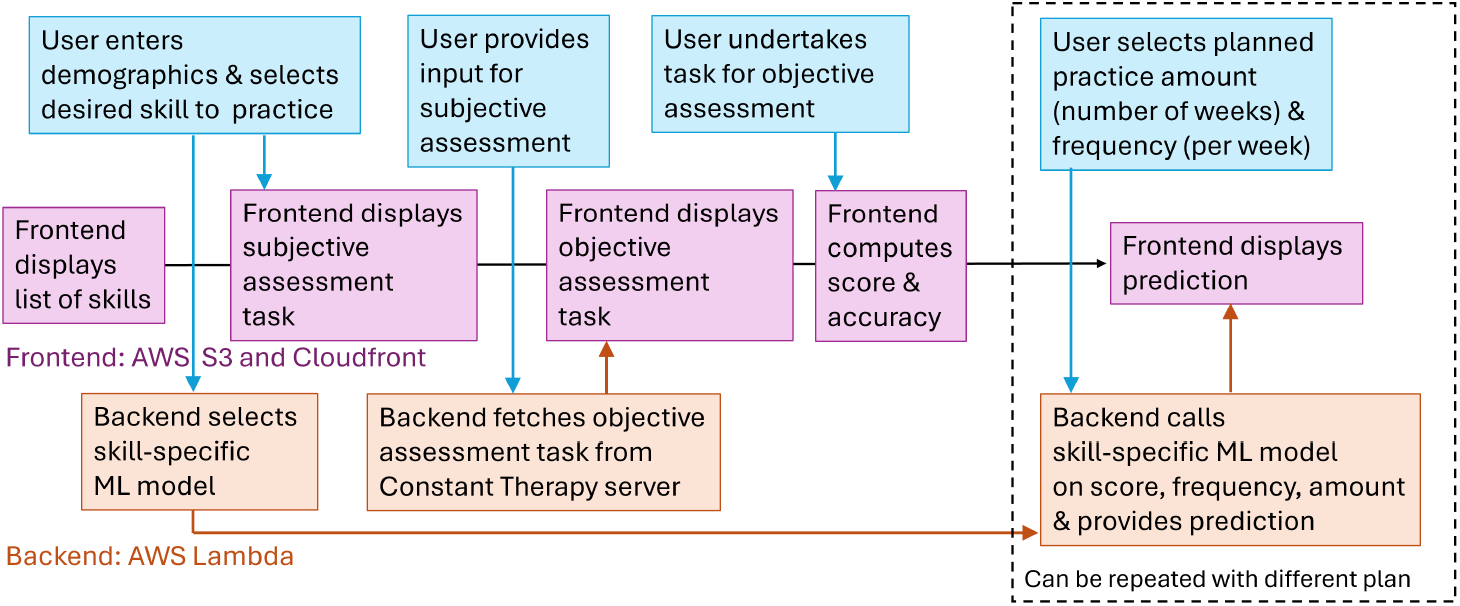
Workflow of the Therapy Calculator with user tasks and frontend and backend of the website. The user can provide updated practice parameters, e.g., to check how much more practice will improve prediction results (dashed box).

Next, the data are sent to the task-specific machine learning model managed by the backend service. It produces predictions based on the user-specified therapy frequency and length, and returns the results to the frontend to display on the user interface. Patients can adjust their desired therapy frequency from once per week to daily and their desired practice length from one week to one year. The frontend service sends any changes in the user’s practice plan to the backend service, which calls the ML model on the updated input values and then returns the prediction results for display.

Sample screenshots of assessment tasks presented by the web interface are shown in Figure 6 and of a prediction result in Figure 1. Importantly, the prediction provided to the user is the probability of improving from the current landmark (based on the initial assessment results) to the subsequent landmark. The probability is shown in the bar with color gradations indicating lower and progressively higher probabilities.

**Figure 6.**
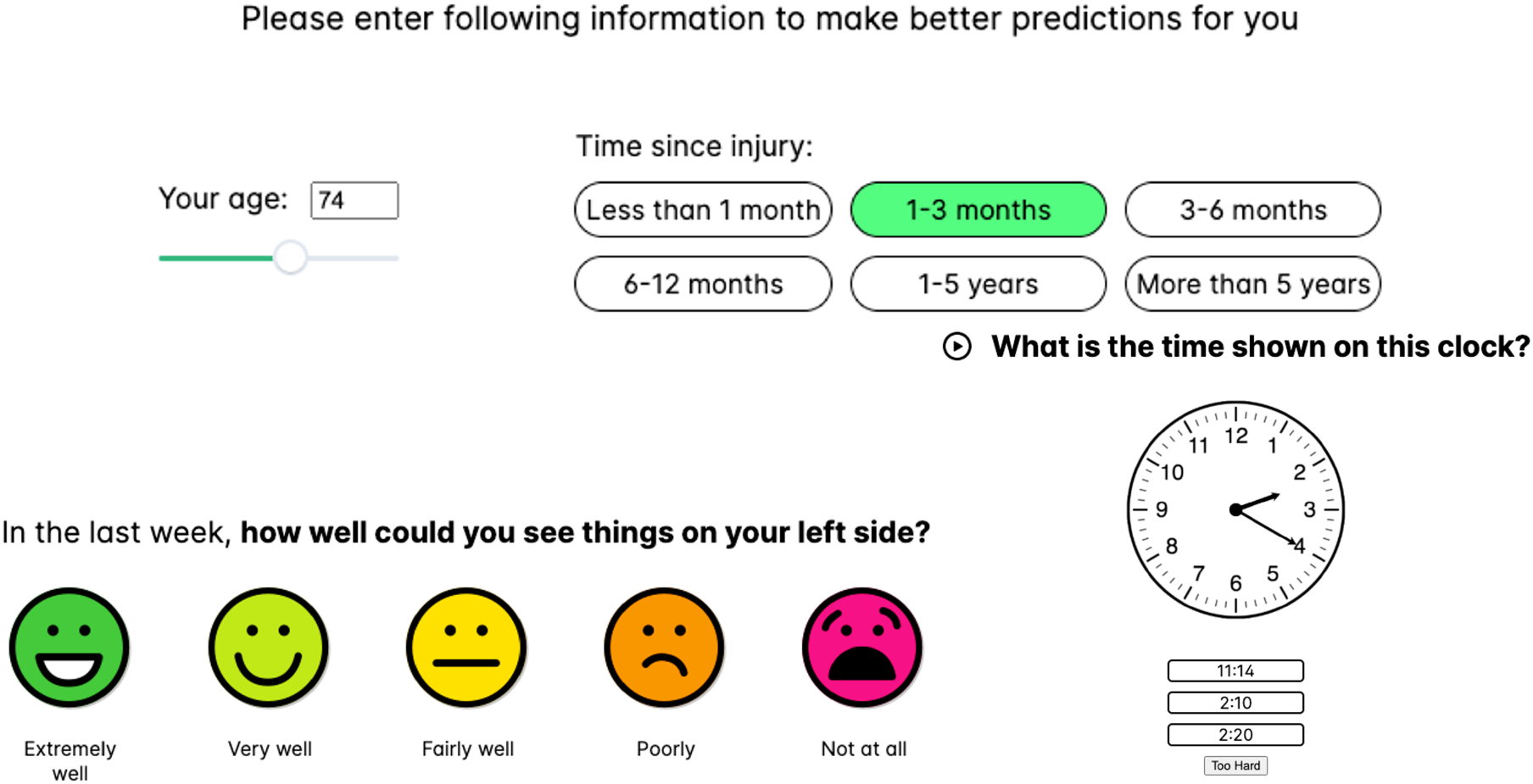
Screenshots of user workflow in the calculator web application. Top: Entering demographic information. Bottom left: a subjective assessment question. Bottom right: an objective assessment task.

Once the Therapy Calculator website was published, we conducted a formative user study. We invited nine stroke patients to test the application and gathered feedback regarding its usability. Feedback provided included (1) addition of audio descriptions for tasks, (2) larger text fonts to improve readability of the interface, and (3) the ability to save and print the calculator results. All these patient suggestions were implemented into the current web interface.

## Discussion

The present study developed an application, called Therapy Calculator, that includes machine learning based prediction and a user-friendly web interface. Therapy Calculator is currently available online (https://www.bu.edu/cbr/research/predictive-modeling/) and has obtained positive feedback from aphasia patients who tested it.

The logistic regression model demonstrated strong predictive performance across all domain-landmark combinations, with particularly high accuracy for lower-numbered landmarks, indicating that the model most reliably predicts patient progress through earlier, foundational therapy tasks. Further, analysis of stroke therapy outcomes revealed several key predictors of patient improvement across cognitive and speech/language domains. In terms of outcomes predictors, younger patients and those treated earlier after stroke achieved better outcomes, while performance metrics such as initial domain scores, and task accuracy emerged as the strongest positive predictors of improvement. Therapy frequency proved highly influential, particularly for domains requiring consistent practice, though total therapy duration showed mixed effects. The influence of these factors varied between cognitive domains, which were more sensitive to demographic features, and speech/language domains, which showed stronger dependencies on initial performance and therapy frequency, particularly at higher skill levels. Notably, consistent practice durations were crucial across all domains, with gaps between sessions negatively impacting therapy progress, especially in areas requiring complex cognitive processing skills.

These results are consistent with the extent literature on the effectiveness of speech, language and cognitive therapy in individuals with post-stroke aphasia^21^. As noted in the introduction, higher dose, higher frequency and intensity (more than 20 to 50 hours in total dosage and 2 to 4 hours per week) of speech-language therapy (SLT) are associated with greater improvements in language and cognitive outcomes^6,22^. Intervention in both the early phases and chronic phases of recovery and functionally tailored therapies that incorporate both receptive and expressive language tasks promote better outcomes^22^. The rapidly growing population of stroke survivors with chronic impairments makes stroke a leading cause of long-term disability, with both societal and personal costs^23–26^. Till date this emerging evidence for the benefits of sustained therapy has not been translated into actionable outcomes for the survivors with aphasia. Indeed, patients do not know whether they will recover or improve their communication and cognitive skills as they live with a long-term disability. While there are similar “calculators” in the stroke/medical field to estimate stroke risk, mortality or functional outcomes (e.g., stroke recovery prediction; ISCORE), this study is the first of its kind therapy outcome calculator to inform patients and clinicians the amount (i.e., dosage, frequency) of therapy required to obtain improvements in speech and cognitive outcomes.

There are several limitations of this work. One limitation of the current model is that it can only predict the likelihood of improvement from one task to the next task in a specific domain. Second, as this study only utilized data for a patient’s initial therapy session and a limited amount of data for further therapy progress (total number of sessions, therapy length, and gap time between sessions), a significant portion of the dataset could not be utilized for model training. Subsequent session data, rich in details like current domain score and accuracy, was omitted as it did not pertain to the model’s input criteria. Further development could involve advanced algorithms, including neural networks, to generate more precise predictions.

## Conclusions

In conclusion, this research has significant implications for personalized therapy planning and outcome prediction in rehabilitation. The development and successful implementation of the Therapy Calculator, (as evidenced by the strong predictive performance) suggests a valuable tool for both clinicians and patients in making data-driven therapy decisions. The key actionable outcomes identify factors that influence recovery outcomes, particularly the importance of early (age and time since injury), consistent practice schedules, and the role of baseline performance in predicting improvement.

The study also showed important distinctions in how different types of therapy domains respond to various factors which can help in tailoring treatment approaches. While the current model has limitations, such as only predicting step-by-step improvement and not utilizing ongoing session data, its positive reception by aphasia patients demonstrates its practical value.

## Author contributions

All authors contributed to methodology design. H.L. performed the experiments and website development. P.I. and M.B. provided help with machine learning framework design and website development. H.L. and S.K. wrote the original draft of the paper while all authors contributed to reviewing the following versions. All authors contributed to the article and approved the submitted version.

## Data availability

The data and code used for this analysis are in the process of being uploaded to Open Science Framework (OSF) and will be available at the following link: http://osf.io/mcu36. In the interim, data are available from the authors upon request.

